# The active lung microbiota landscape of COVID-19 patients

**DOI:** 10.1101/2020.08.20.20144014

**Authors:** Yang Han, Zhilong Jia, Jinlong Shi, Weidong Wang, Kunlun He

## Abstract

With the outbreak of COVID-19 causing by SARS-CoV-2, the interaction between the host and SARS-CoV-2 was widely studied. However, it is unclear whether and how SARS-CoV-2 infection affects lung microflora, which contributes to COVID-19 complications. Here, we analyzed the metatranscriptomic data of bronchoalveolar lavage fluid (BALF) of 19 COVID-19 patients and 23 healthy controls from 6 independent projects and detailed the active microbiota landscape in both healthy individuals and COVID-19 patients. The infection of SARS-CoV-2 could deeply change the lung microbiota, evidenced by the α-diversity, β-diversity and species composition analysis based on bacterial microbiota and virome. Pathogens (such as *Klebsiella oxytoca* causing pneumonia as well), immunomodulatory probiotics (such as Lactic Acid Bacteria and *Faecalibacterium prausnitzii*, a butyrate producer) and Tobacco mosaic virus (TMV) were enriched in the COVID-19 group, suggesting a severe microbiota dysbiosis. The significant correlation between *Rothia mucilaginosa*, TMV and SARS-CoV-2 revealed drastic inflammatory battles between the host, SARS-CoV-2 and other microbes in the lungs. Notably, TMV only existed in the COVID-19 group, while Human respirovirus 3 only existed in the healthy group. Our study provides insight into the active microbiota in the lungs of COVID-19 patients and will contribute to the understanding of the infection mechanism of SARS-CoV-2 and the treatment of the disease and complications.

## Introduction

With the outbreak of severe acute respiratory syndrome coronavirus 2, SARS-CoV-2(1), leading to Coronavirus disease 2019 (COVID-19), many studies on clinical characteristics, three-dimensional structure(2) and interactions between host and virus(3) were reported. The symptoms of COVID-19 patients (COV) usually include fever, vomiting, diarrhea(4). As bacterial or viral infections could also induce several symptoms among them, it is necessary to profile the microbiota of COVID-19 patients to effectively relieve their symptoms(5, 6). The bronchoalveolar lavage fluid (BALF), containing microenvironment information on bronchioles and lung alveoli from the lower respiratory tract, is one of key sample types for characterizing the host inflammatory response and microbiota of COVID-19 patients as lung is one of main organs for the infection of SARS-CoV-2(7, 8).

The human microbiota in BALF, including bacterial microbiota and virome, is a diverse microbial ecosystem associated with beneficial or deleterious physiological functions as well as disease etiologies, including COVID-19(9). It was reported that multiple common respiratory pathogens were co-infected with SARS-CoV-2 in COVID-19 patients(10). Moreover, the role of commensal viruses in human lungs is poorly understood, particularly in the respiratory tract(11). Besides, there are close interactions between microorganisms including symbiosis and competition(12, 13). Therefore, the relationship between SARS-CoV-2 and other microorganisms in the body is of great significance for studying its infection mechanism and developing effective treatments. Shen et al. briefly reported that the bacterial microbiota in COVID-19 patients was dominated by the pathogens and oral and upper respiratory commensal bacteria(14). However, there is no systematic microbiota landscape in BALF samples from COVID-19 patients as well as the healthy individuals.

In this study, we systematically profiled the transcriptionally active microbiota landscape in BALF from COVID-19 patients and healthy individuals, identified microorganism composition in healthy individuals and COVID-19 patients, found disease-specific active microbes in the COVID-19 patient group, revealed the interaction between several bacteria or viruses and SARS-CoV-2. The systematic microbiota landscape provided crucial insight into understanding the mechanism of SARS-CoV-2 infection and the treatment of COVID-19.

## Materials and Methods

### Data collection and preprocessing

The raw metatranscriptomic fastq data of for 42 BALF samples, consist of 19 COVID-19 patients and 23 healthy controls, and two negative controls (NC) were downloaded from Sequence Read Archive, EMBL-EBI ArrayExpress and National Genomics Data Center, China with project IDs, PRJCA002202, PRJNA601736, PRJCA002326, PRJNA605983, PRJNA615032 and PRJNA434133. The basic information of these samples was described in Supplementary Table 1. The raw fastq data were analyzed using IDSeq webserver(15) (Pipeline v4.9) to obtain the raw read count of each bacterial and viral species. Briefly, the raw reads underwent removal of host reads (Hg38 reference genome) via STAR (Spliced Transcripts Alignment to a Reference), a series of quality control, consist of removal of low-quality, low-complexity and redundant sequences via Paired-Read Iterative Contig Extension (PRICE), CD-HIT-DUP and Lempel-Ziv-Welch (LZW) compression score, respectively(15). After quality control, the assignment of taxonomic IDs was implemented by aligning to the NCBI nucleotide (nt) and non-redundant protein (nr) databases via GSNAPL and RAPsearch(15), respectively. Then the read counts of nt and reads per million (rPM) of nt at the species level were obtained. To guarantee the quality and credibility of the lung microbiota due to the low biomass, species with rPM value less than 50 in NC were used for downstream analysis. Moreover, bacteria detected in at least two study projects were kept to largely eliminate the reagent and laboratory contamination.

### Microbial diversity and differential analysis

Microbial diversity was analyzed by R package vegan (v2.5-6). Shannon diversity index was calculated to evaluate α-diversity. Bacterial or viral loads were computed based on rPM of microorganism. β-diversity was studied by Principal coordinate analysis, PCoA, using R package ape (v5.3) based on Bray-Curtis distance. Differential species were analyzed by DEseq2 (v1.24.0). Differential bacteria with a statistical significance (q-value) < 1 e-11 and absolute value of log_2_(Fold Change) > 3, virus with a statistical significance (q-value) < 0.01 and absolute value of log_2_(Fold Change) > 1 were retained. Student’s t test and Mann-Whitney rank test were used in variables with normal and without normal distribution, respectively. The Benjamini-Hochberg correction was used to obtain FDR adjusted p-values (q-values) for multiple hypothesis testing.

### Microbiota correlation analysis in BALF

Correlation analysis was implemented by Spearman’s rank correlation. Only bacteria and viruses retained in the differential analysis were used for correlation analysis. Bacteria correlations with a coefficient (r) > 0.8 and a statistical significance (q-value) < 1 e-13 were considered for further analysis and visualization. Virus correlations with a coefficient (r) > 0.5 and a statistical significance (q-value) < 0.01 were considered for further analysis and visualization. Bacteria and virus correlations with a coefficient (r) > 0.7 and a statistical significance (q-value) < 5e-8 were considered for further analysis and visualization. Correlation network was visualized by Cytoscape 3.7.2(16).

## Results

We collected the metatranscriptomic data of 42 BALF samples, consist of 19 COVID-19 patients and 23 healthy controls (HC), from six studies(1, 14, 17-20). The basic information of the samples was shown in Supplementary table 1. 19 COVID-19 patients were from five different studies, representing individual sample source of the samples. In totally, 12413 bacteria and 359 viruses at the species level, which were transcriptionally active, were identified.

### Differential microbial diversity in the COVID-19 and healthy groups

The diversity analysis of bacteria and virus, including α-and β-diversity, showed large differences between two groups. The COVID-19 group had higher Shannon’s Diversity Index of bacteria than healthy group (p-value = 0.0268, Student,s t test, Figure 1a). Also, Bacterial loads of COVID-19 group were much higher than healthy group (p-value = 9e-4, Mann-Whitney rank test, Figure 1b). For virome, COVID-19 patients had much higher viral loads than healthy controls (p-value < 1e-4, Mann-Whitney rank test) while Shannon’s Diversity Index showed no statistical difference (Figure 1cd).

**Figure 1.**
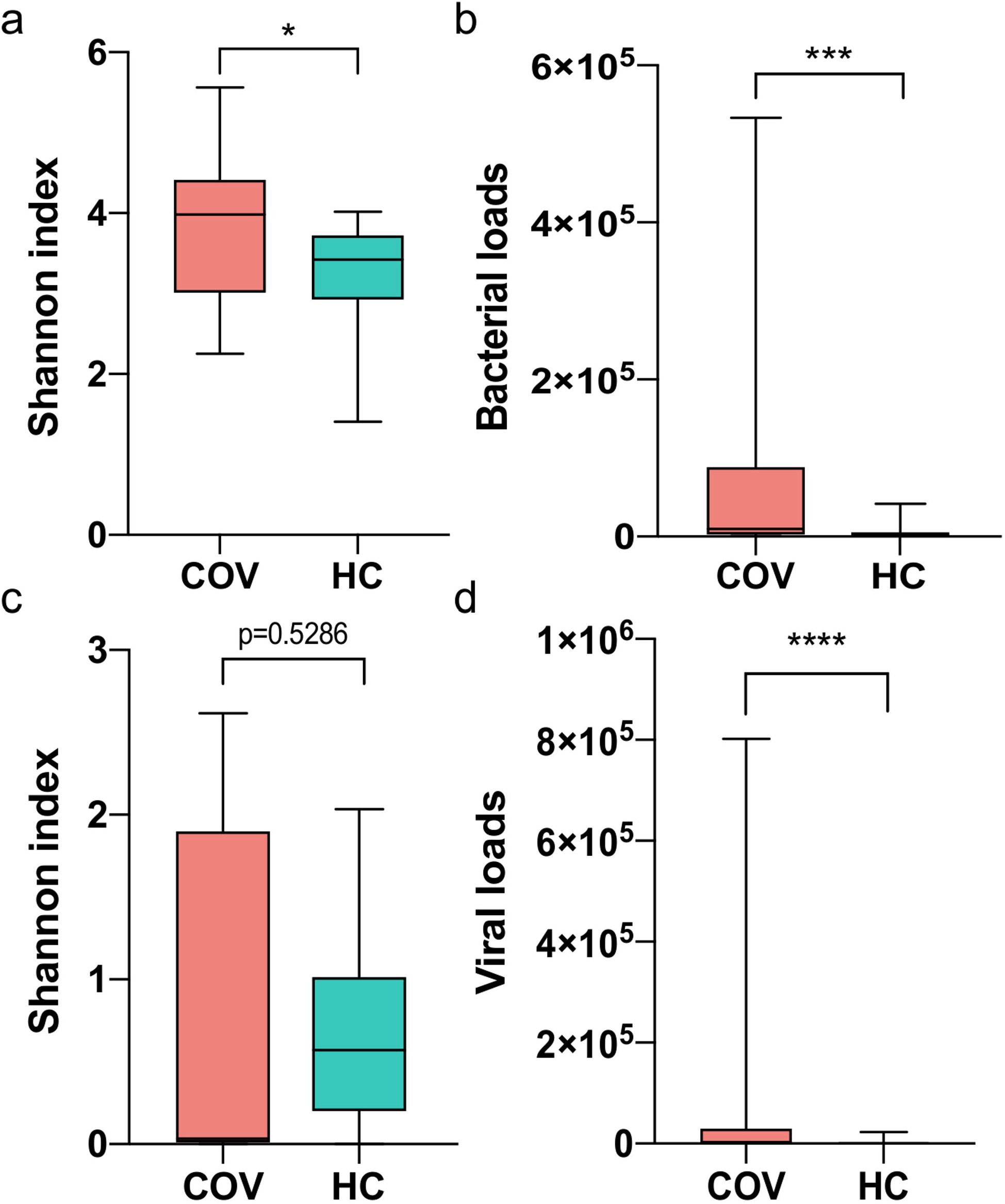
Comparison of microbial α-diversity and loads between COVID-19 patients (COV) and healthy controls (HC) (a) and (c) Shannon index of bacteria and virus respectively. (b) and (d) Bacterial and viral loads respectively. Boxes represent the interquartile ranges (IQRs) between the first and third quartiles, and the line inside the box represents the median; whiskers represent the lowest or highest values within 1.5 times IQR from the first or third quartiles. * p-value < 0.05, ** p-value < 0.01, *** p-value <0.001, **** p-value <0.0001.

Furthermore, Principal Coordinates Analysis (PCoA) with Bray-Curtis distance was applied to evaluate the β-diversity in BALF microbiota across groups. The PCoA results of bacterial microbiome showed there was a significant difference of β-diversity between two groups (p-value < 1e-3, R = 0.217, ANOSIM Analysis) (Figure 2a). Also, similar results for virome were found (p-value < 1e-3, R = 0.541, ANOSIM Analysis) (Figure 2b), indicating a heterogeneous community diversity in the patient and control groups. The diversity analysis revealed that the infection of SARS-CoV-2 probably caused a different lung microbiota composition in the COVID-19 patient group compared with the healthy group.

**Figure 2.**
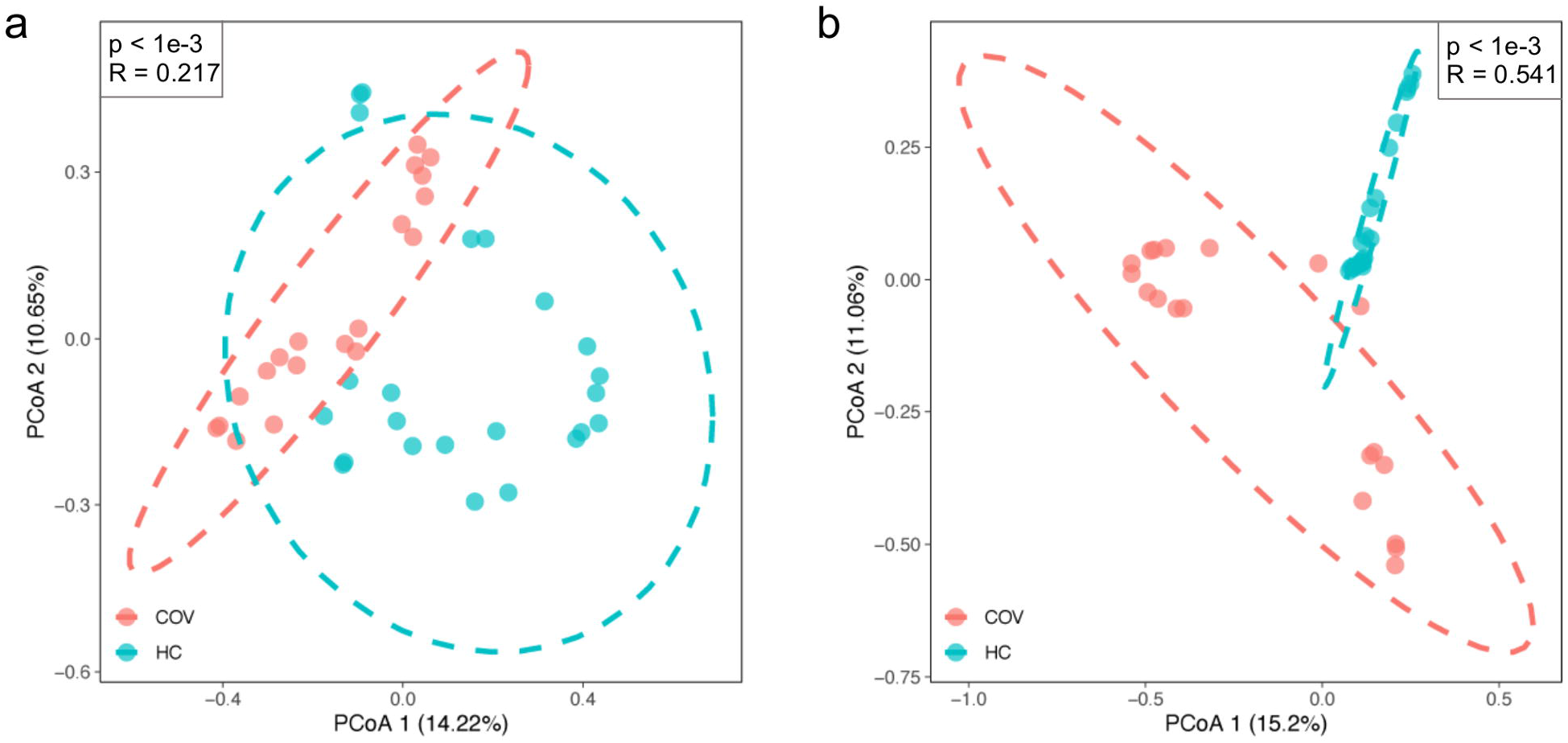
β-diversity of bacteria and viruses between COVID-19 patients (COV) and healthy controls (HC) (a) and (b) Principal coordinate analysis plot of bacteria and virus respectively based on Bray-Curtis distance. The pink dot indicates COVID-19 patients and the light green indicates healthy controls. R and p-value are the results of ANOSIM test.

### Microbial composition of BALF samples

To get an insight of the microbial composition of BALF, we screened out the 10 most abundant bacteria at the species level in healthy controls and COVID-19 patients (Figure 3a), respectively. Concerning the bacteria community, *Porphyromonas gingivalis*, often found in the oral cavity where it is implicated in periodontal disease(21), had a relatively higher abundance in some healthy individuals. The abundance of *Lautropia mirabilis*, which was commonly found in the human oral cavity and the upper respiratory tract(22), and *Enterobacter kobei*, leading to nosocomial infections(23), were increased in a few COVID-19 patients. Notably, *Variovorax* account for a very high proportion in 6 patients. Interestingly, *Staphylococcus xylosus* and *Staphylococcus simulans* had high abundances in three healthy controls (H21, H22 and H23).

**Figure 3.**
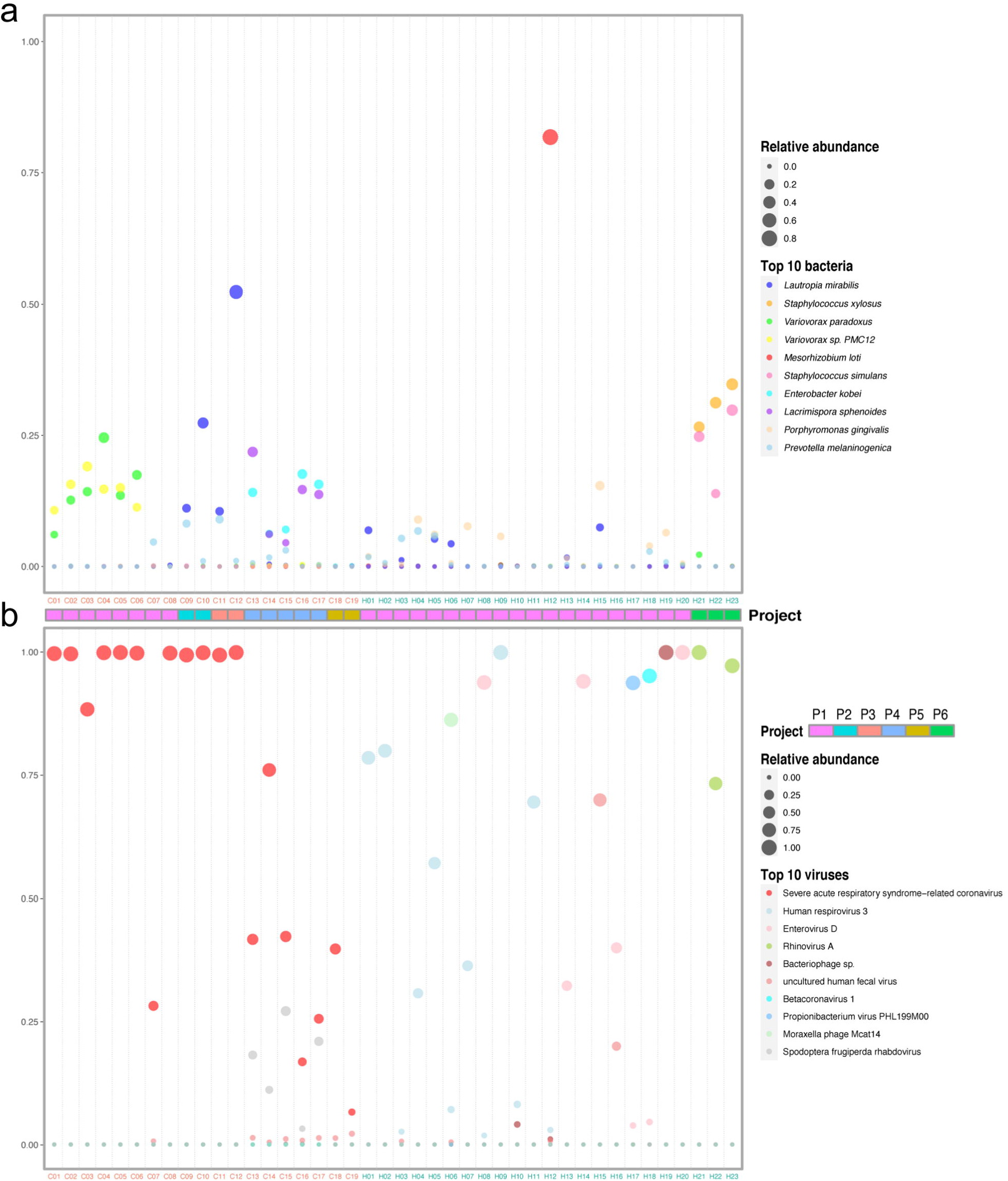
Composition of the top 10 species in COVID-19 patients and healthy controls. (a) and (b) Relative abundance of the top 10 bacteria and viruses respectively in each sample. The size of the point corresponds to the relative abundance value. The color of the sample names represents the sample group, red represents the COVID-19 patient group, and blue represents the healthy control group. The order of the species in the legend from top to bottom represents the relative abundance from high to low. Project bar represents the sample sources (P1, PRJCA002202; P2, PRJNA601736; P3, PRJCA002326; P4, PRJNA605983; P5, PRJNA615032; P6, PRJNA434133).

The virome composition analysis showed that Human respirovirus 3 and Enterovirus D were higher in a few healthy samples (Figure 3b). Except severe acute respiratory syndrome-related coronavirus, Spodoptera frugiperda rhabdovirus was significantly enriched in five patient samples. (Figure 3b). Severe acute respiratory syndrome-related coronavirus includes Severe acute respiratory syndrome coronaviruses (SARS-CoVs) and SARS-CoV-2. The top abundant species showed that the infection of SARS-CoV-2 changed the bacterial and viral community in the lung of COVID-19 patients.

### Microbial differences between the COVID-19 patient and healthy control groups

To identify the species associated with COVID-19, differential abundance analysis was performed between the two groups. 99 bacteria at the species level were significantly different between the two groups (q-value < 1e-11, absolute value of log_2_(Fold Change) > 3) (Supplementary Table 2). Meanwhile, 82 species with the maximum relative abundance greater than 0.05% were selected from the 99 bacteria and visualized using heatmap to get a deeper investigation (Figure 4a).

**Figure 4.**
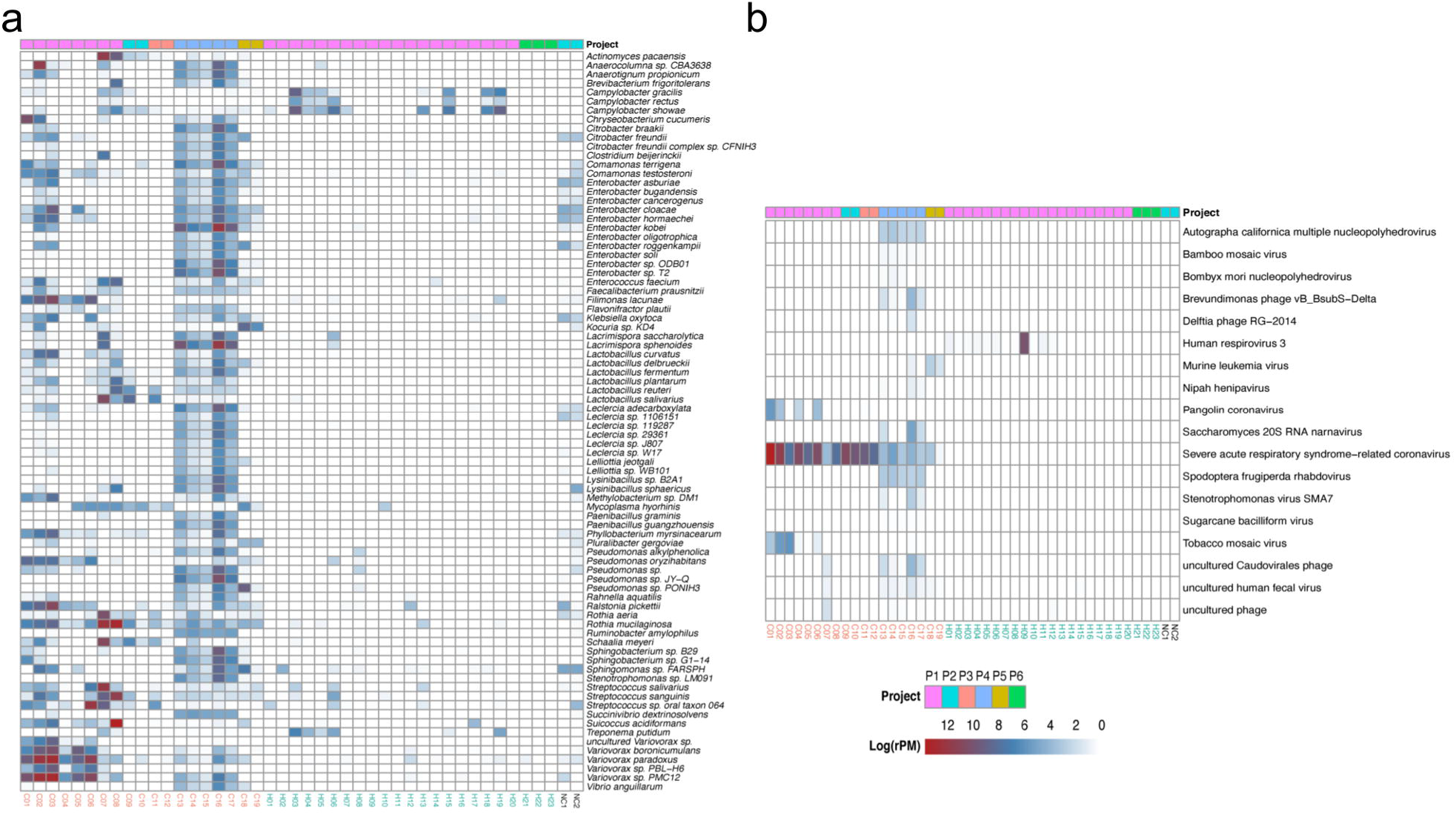
Heat map of the species with strongly significant differences and important differential pathogens. (a) Heat map of differential bacteria with q-value < 1e-11, absolute value of log_2_(Fold Change) >3 and the maximum relative abundance greater than 0.05%. (b) Heat map of virus with q-value < 0.01 and absolute value of log_2_(Fold Change) >1. Project bar represents the sample sources (P1, PRJCA002202; P2, PRJNA601736; P3, PRJCA002326; P4, PRJNA605983; P5, PRJNA615032; P6, PRJNA434133). The color of the sample names represents the sample group, red represents the COVID-19 patient group, and blue represents the healthy control group. NC1 and NC2 are negative controls.

The abundance of Lactic Acid Bacteria (LAB), such as *Lactobacillus fermentum, Lactobacillus reuteri, Lactobacillus delbrueckii* and *Lactobacillus salivarius*, were higher in the COVID-19 group than in the healthy group (Supplementary Table 2). Of note, *Lactobacillus reuteri* was detected only in the patient group. Some pathogens were enriched in COVID-19 patients and depleted in healthy controls. For example, *Klebsiella oxytoca*, leading to pneumonia, colitis and sepsis(24), and *Enterobacter cloacae*, positively correlated with COVID-19 severity(25), were increased in COVID-19. The abundance of some nosocomial infection pathogens, such as *Enterobacter* kobei(23), *Enterobacter cloacae(26)* and *Ralstonia pickettii(27)*, were higher in patients than healthy controls. Several gut bacteria like *Faecalibacterium prausnitzii(28), Enterococcus faecium(29)* and *Citrobacter freundii(30)* and commensal bacteria residing in the mouth and respiratory tracts, such as *Rothia mucilaginosa*, were also enriched in the lung of COVID-19 patients. In addition, a few of pathogens causing severe nausea, vomiting and diarrhea, like *Bacillus* cereus(31) increased in patients significantly.

The abundance of 18 viruses significantly differed between the two groups (q-value < 0.01, absolute value of log_2_(Fold Change) > 1) (Figure 4b, Supplementary Table 3). In addition to severe acute respiratory syndrome-related coronavirus, Surprisingly, 13 of 18 differential viruses were only detected in the COVID-19 group, such as Pangolin coronavirus and Tobacco mosaic virus (TMV), which were probably closely related to the infection of SARS-CoV-2, though the function of them is unclear. Notably, Human respirovirus 3 was only detected in healthy controls (12 of 23 individuals).

### Microbiota correlation in BALF

To further investigate whether the altered microbiota interacted with each other, we constructed networks of bacteria and viruses based on Spearman rank correlations, respectively. Network within bacteria (Figure 5 a and Supplementary Table 4) showed that lots of bacteria were highly correlated. For example, *Enterobacter sp., Pseudomonas sp*. and *Leclercia sp*. were positively correlated to each other. What’s more, *Anaerotignum propionicum*, detected only in the patient group, was positively correlated to *Chryseobacterium cucumeris, Ruminococcus gnavus* and *Leclercia sp. LSNIH1*.

**Figure 5.**
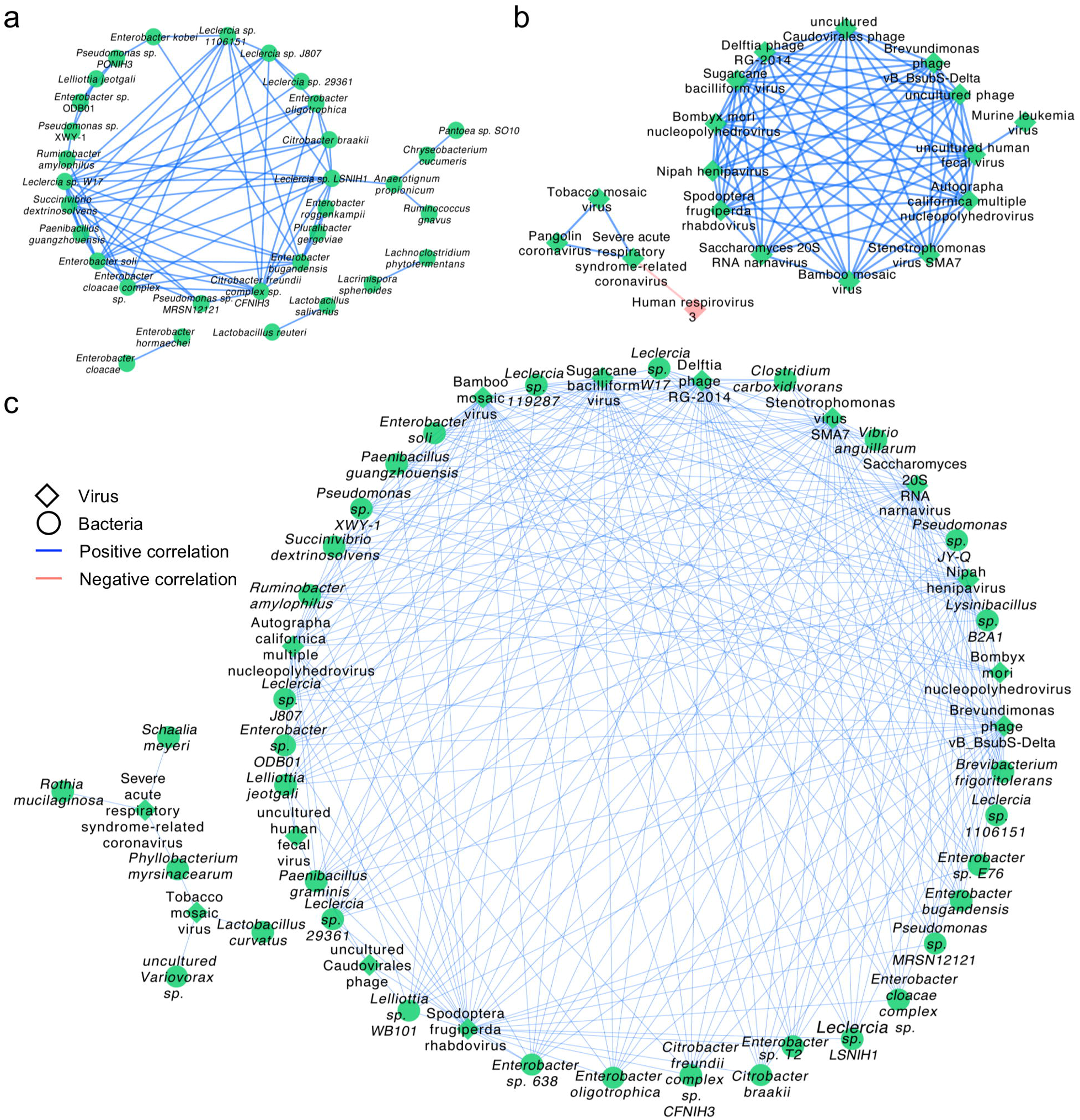
Correlation network of differential species between the COVID-19 patient and healthy control groups. (a) The correlation network within bacteria with a coefficient (r) > 0.8 and a statistical significance (q-value) < 1e-13. (b) The correlation network within viruses with a coefficient (r) > 0.5 and a statistical significance (q-value) < 0.01. (c) The correlation between bacteria and viruses with a coefficient (r) > 0.7 and a statistical significance (q-value) < 5e-8. Diamonds represent viruses, circles represent bacteria. Blue line represents positive correlations and pink line represents negative correlations. Green represents up-regulation of the abundance of the species and pink represents down-regulation.

Network within viruses (Figure 5b and Supplementary Table 5) showed severe acute respiratory syndrome-related coronavirus, TMV and Pangolin coronavirus were positively correlated to each other, while severe acute respiratory syndrome-related coronavirus was negatively correlated to Human respirovirus 3. The Network between bacteria and viruses (Figure 5c and Supplementary Table 6) presented that severe acute respiratory syndrome-related coronavirus was positively correlated to *Rothia mucilaginosa, Schaalia meyeri* and *Phyllobacterium myrsinacearum* which was also positively related to TMV.

Taken together, the results of correlation analysis suggest that the altered lung microbiota, particularly species from the *Lactobacillus* and *Rothia*, *Enterobacter* and TMV, actively played a certain role during the infection of SARS-CoV-2. The negative correlation between SARS-CoV-2 and Human respirovirus 3 indicates that the former probably have an inhibitory effect on the latter.

## Discussion

The transcriptionally active microbiota landscape of BALF in COVID-19 patients and healthy controls could shed light on the infection mechanism of SARS-COV-2. Our study described the microbial composition at the species level in BALF of COVID-19 patients and healthy controls. Although *Porphyromonas gingivalis* existed widely in healthy lungs and accounted for a high proportion, lower richness and bacterial loads means it still stay a low level. In addition, though Human respirovirus 3 is one of the most common respiratory viruses, particularly among young children under 5 years of age and immunocompromised patients(32, 33), it was detected in the healthy group with no symptoms.

The infection of SARS-CoV-2 changed the active microbiota in the lungs, characterized by the α-diversity, β-diversity, species composition of bacteria and viruses respectively. The changes should be a result of complex interactions between the host, SARS-CoV-2, bacteria and other viruses. The enrichment of LAB, probiotics usually producing lactic acid, in the COVID-19 group could probably inhibit the inflammatory response caused by SARS-CoV-2 to some extent, as several species of *Lactobacillus* could decrease the inflammatory response, and balance the immune response while limiting the potential of pathogens(34, 35). Besides, *Lactobacillus* could help treat diarrhea. The enrichment of several pathogens, such as bacteria *(Klebsiella oxytoca, Enterobacter kobei* and *Bacillus cereus)* and viruses (Pangolin coronavirus and TMV), in the COVID-19 group probably indicated that co-infections or a second infection. Several symptoms, such as gastrointestinal symptoms and extreme fatigue in addition to fever, may be caused by the increase of these pathogens.

Although *Rothia mucilaginosa* was a part of the normal oropharyngeal flora, it was linked to causing disease, including pneumonia, in immunosuppressed humans(36). The enrichment of it in COVID-19 patients may be the result of the impact of SARS-CoV-2 on the patients’ immune system. In our study, we also found that some intestinal bacteria (such as *Faecalibacterium prausnitzii* and *Enterococcus faecium)* were enriched in the lung of COVID-19 patients. A study reported that intestinal bacteria could transfer to other organs through the intestinal barrier(37), so we speculated that the infection of SARS-CoV-2 could probably promote the transfer of intestinal bacteria. *Faecalibacterium prausnitzii*, a prototypical IL-10 and butyrate producer in the gut(38), showed a negative correlation with COVID-19 severity(25). Therefore, it has the potential to be a protector of the host for the infection.

Subjects with COVID-19 have unexpected colonization of the lung by TMV and Pangolin coronavirus, suggesting that they may play a role in the infection of SARS-CoV-2. It was reported that TMV could enter and persist in mouse lungs, raising questions about the potential interactions between the virus and human host(39). Accordingly, a further investigation was necessary. Our BALF metatranscriptome results detailed the species in the lungs of COVID-19 patients and shed light on the complex interactions between SARS-CoV-2 and other microbes in the lungs.

## Conclusion

The metatranscriptome of human BALF can reveal transcriptionally active microbiota in the lungs. The microbes in the lungs have a great influence on human health and disease. We detailed the active microbiota landscape in both healthy individuals and COVID-19 patients and found the infection of SARS-CoV-2 could deeply change the microbiota in the lungs. This is the first detail description of lung colonization in COVID-19 patients. The enrichment of several microbes, Lactic Acid Bacteria, *Faecalibacterium prausnitzii* and TMV, the significant correlation between *Rothia*, TMV and SARS-CoV-2 reveals a microbiota disorder and drastic battles between the host, SARS-CoV-2 and other microbes in the lungs. Our study provides insight into the active microbiota in the lungs of COVID-19 patients and will contribute to the understanding of the infection mechanism of SARS-CoV-2 and the treatment of the disease and complications.

## Data Availability

The raw datasets analyzed for this study were downloaded from Sequence Read Archive, EMBL-EBI ArrayExpress and National Genomics Data Center, China with project IDs, PRJCA002202, PRJNA601736, PRJCA002326, PRJNA605983, PRJNA615032 and PRJNA434133.

## Authors’ Contributions

Conceptualization: Z. J., K.H. and W.W.; Data analysis: Y.H., Z.J., J.S.; Writing & Editing: Y.H. and Z.J. All authors read and approved the final manuscript.

## Supplementary tables

Supplementary table 1. The basic information of the samples. The table lists the Project ID, sample ID, sample name, total reads, non-host reads, the percentage of non-host reads, water control and collection location.

Supplementary table 2. Differential bacteria at the species level (q-value < 1 e-11, absolute value of log_2_(Fold Change) > 3).

Supplementary table 3. Differential viruses at the species level (q-value < 0.01, absolute value of log_2_(Fold Change) > 1).

Supplementary table 4. Correlation within differential bacteria (r > 0.8, q-value < 1e-13).

Supplementary table 5. Correlation within differential viruses (r > 0.5, q-value < 0.01).

Supplementary table 6. Correlation of differential bacteria with differential viruses (r > 0.7, q-value < 5e-8).

